# Comparative evaluation of six immunoassays for the detection of antibodies against SARS-CoV-2

**DOI:** 10.1101/2020.09.08.20190488

**Authors:** Felipe Pérez-García, Ramón Pérez-Tanoira, María Esther Iglesias, Juan Romanyk, Teresa Arroyo, Peña Gómez-Herruz, Rosa González, Juan Cuadros-González

## Abstract

**Objectives:** Serologic techniques can serve as a complement to diagnose SARS-CoV-2 infection. The objective of our study was to compare the diagnostic performance of six immunoassays to detect antibodies against SARS-CoV-2: three lateral flow immunoassays (LFAs), one ELISA and two chemiluminescence assays (CLIAs).

**Methods:** We evaluated three LFAs (*Alltest, One Step and SeroFlash*), one ELISA (*Dia.Pro*) and 34 two CLIAs (*Elecsys* and *COV2T*). To assess the specificity, 60 pre-pandemic sera were 35 used. To evaluate the sensitivity, we used 80 serum samples from patients with 36 positive PCR for SARS-CoV-2. Agreement between techniques was evaluated using the kappa score (*k*).

**Results:** All immunoassays showed a specificity of 100% except for *SeroFlash* (96.7%). Overall sensitivity was 61.3%, 73.8%, 67.5%, 85.9%, 88.0% and 92.0% for *Alltest, One Step, SeroFlash, Dia.Pro, Elecsys* and *COV2T*, respectively. Sensitivity increased throughout the first two weeks from the onset of symptoms, reaching sensitivities over 85% from 14 days for all LFAs, being *One Step* the most sensitive (97.6%), followed by *SeroFlash* (95.1%). *Dia.Pro, Elecsys* and *COV2T* showed sensitivities over 97% from 14 days, being 100% for *COV2T*. *One Step* showed the best agreement results among LFAs, showing excellent agreement with *Dia.Pro* (agreement=94.2%, *k*=0.884), *COV2T* (99.1%, *k*=0.981) and *Elecsys* (97.3%, *k*=0.943). *Dia.Pro, COV2T* and *Elecsys* also showed excellent agreement between them.

**Conclusion:** *One Step, Dia.Pro, Elecsys* and *COV2T* obtained the best diagnostic performance results. All these techniques showed a specificity of 100% and sensitivities over 97% from 14 days after the onset of symptoms, as well as excellent levels of agreement.

## 1. Introduction

Since the beginning of the pandemic due to SARS-CoV-2 in Wuhan in December 2019, the virus has caused as of August 19, 2020, more than 21 million cases and 775,000 deaths worldwide [1]. An early and accurate diagnosis of SARS-CoV-2 infection is essential for the adequate management of COVID-19 patients and the establishment of infection control measures in order to contain this pandemic. Polymerase chain reaction (PCR) is the reference method for COVID-19 diagnosis but its sensitivity depends on the type of sample (upper or lower respiratory tract) and the time from infection [2–4]. Serologic tests are useful tools for the diagnosis and management of SARS-CoV-2 infection as a complement of PCR [5]. They have also proved their usefulness in epidemiological surveillance studies [6,7]. The vast majority of these tests are based on the detection of IgM and IgG antibodies against SARS-CoV-2 nucleocapsid and spike [8]. There are mainly three diagnostic approaches to detect these antibodies: lateral flow immunoassays (LFAs), enzyme-linked immunoassays (ELISAs) and chemiluminescence immunoassays (CLIAs). The aim of our study was to compare the diagnostic performance of six serologic tests for the detection of antibodies against SARS-CoV-2: three LFAs, one ELISA and two CLIAs.

## 2. Methods

### 2.1. Population and study period

The study was performed between 1st March and 30th April 2020. We included two groups of patients:

<u>Negative controls</u>: 60 serum samples from a randomly selected group of patients who the sample taken for other serologic studies, from September 1 to November 30, 2019. Aliquots of cryopreserved sera were recovered from the serum archive.

<u>PCR positive patients:</u> 80 patients admitted to the Emergency department between March 1 and April 28, 2020, with suspicion of COVID-19 and confirmation by PCR. All of them were symptomatic, with a median time from the onset of symptoms of 15 days (Interquartile range, 8 – 25 days). Residual serum samples were recovered for this evaluation.

### 2.2. Serological assays

<u>LFAs:</u> we evaluated one LFA which detects IgG and IgM antibodies against SARS-CoV-2 nucleocapsid (*AllTest COVID-19 IgG/IgM* [AllTest Biotech, Hangzhou, China]) and two LFAs which detect IgG and IgM antibodies against nucleocapsid and spike ( *One Step Rapid Test* [Innovita Biological Technology, Hebei, China] and *SeroFlash SARS-CoV-2 IgM/IgG* [Epigentek Group, New York, USA]).

<u>ELISA:</u> we evaluated one ELISA which detects IgG and IgM antibodies against nucleocapsid and spike (*Dia.Pro COVID-19* [Dia.Pro Diagnostic Bioprobes, Sesto San Giovanni, Italy]).

<u>CLIA:</u> we evaluated two CLIAs for total antibodies (IgM+IgG): *Elecsys Anti-SARS-CoV-2* (Roche Diagnostics, Mannheim, Germany), which detects antibodies against nucleocapsid and *SARS-CoV-2 Total COV2T* (Siemens Healthineers, Erlangen, Germany), which detects antibodies against spike (S1). Sensitivity evaluation of CLIA techniques could only be performed with 50 samples due to insufficient sample volume.

### 2.3. Statistical analysis

We considered a positive result for samples in which IgG, IgM or both of them were detected. Specificity and sensitivity with 95% confidence intervals (95%CI) were calculated using the results from negative controls and positive PCR patients, respectively. Sensitivity was evaluated globally and also according to the time from the onset of symptoms. Agreement between different techniques was evaluated using the Cohen’s *kappa* score [9]. Statistical analysis was performed using Stata/IC 13.1 (StataCorp, Texas, USA).

## 3. Results

Overall serologic results from the different techniques are summarized in **Table 1**. All techniques showed a specificity of 100%, except for *SeroFlash* LFA, which showed a specificity of 96.7% due to two samples that were positive for IgM antibodies. Regarding LFAs, the overall sensitivity was 61.3% for *Alltest*, 73.8% for *One Step* and 67.5% for *SeroFlash*. *Dia.Pro* ELISA showed an overall sensitivity of 85.9% and CLIAs showed sensitivities of 92.0% for *COV2T* and 88.0% for *Elecsys*. Regarding LFAs, sensitivity increased within the first two weeks (**Table 2**), reaching a sensitivity over 85% from 14 days from the onset of symptoms for all of them. The most sensitive LFA was *One Step* (96.7%), followed by *SeroFlash* (95.1%). ELISA and CLIAs showed sensitivities over 97% from 14 days from the onset of symptoms, being 100% for *COV2T* CLIA (**Table 2**). Finally, the results for agreement between techniques are summarized in **Table 3**. *One Step* showed the best agreement results among the evaluated LFAs, showing almost perfect agreement with *SeroFlash* (agreement = 92.1%, *k* = 0.838), *Dia.Pro* (94.2%, *k* = 0.884), *COV2T* (99.1%, *k* = 0.981) and *Elecsys* (97.3%, *k* = 0.943). *Dia.Pro* ELISA showed almost perfect agreement with CLIAs (98.2%, *k* = 0.963 for *COV2T*; 96.4%, *k* = 0.925 for *Elecsys*). Finally, *COV2T* and *Elecsys* CLIAs also showed almost perfect agreement between them (98.2%, k = 0.962).

**Table 1.** Overall diagnostic performance of the evaluated immunoassays

| Serologic test | PCR positive patients | Negative controls | Sensitivity (95%CI) | Specificity (95%CI) |
| --- | --- | --- | --- | --- |
| **LFA** |  |  |  |  |
| *Alltest (Alltest)* | 49/80 | 0/60 | 61.3<br>(49.7 – 71.9) | 100.0<br>(94.0 – 100.0) |
| *One Step (Innovita)* | 59/80 | 0/60 | 73.8<br>(62.7 – 83.0) | 100.0<br>(94.0 – 100.0) |
| *Seroflash (Epigentek)* | 54/80 | 2/60 | 67.5<br>(56.1 – 77.6) | 96.7<br>(88.5 – 99.6) |
| **ELISA** |  |  |  |  |
| *Dia.Pro (Dia.Pro)* | 67/78* | 0/60 | 85.9<br>(76.2 – 92.7) | 100.0<br>(94.0 – 100.0) |
| **CLIA** |  |  |  |  |
| *COV2T (Siemens)* | 46/50** | 0/60 | 92.0<br>(80.8 – 97.8) | 100.0<br>(94.0 – 100.0) |
| *Elecsys (Roche)* | 44/50** | 0/60 | 88.0<br>(75.7 – 95.5) | 100.0<br>(94.0 – 100.0) |

**Statistics:** values are expressed as absolute count (percentage). A positive serologic result was defined for LFA and ELISA tests for samples that resulted positive for either IgM or IgG antibodies. **Abbreviations:** LFA: lateral flow assay; ELISA: enzyme-linked immunoassay; CLIA: chemiluminescence; 95%CI: 95% confidence interval.
\* Two samples presented indeterminate result for IgG or IgM and were excluded from the analysis
\*\* Sensitivity evaluation of CLIA techniques could only be performed with 50 samples due to insufficient sample volume

**Table 2.** Sensitivity results according to the time from the onset of symptoms

| Serologic test | 0 – 7 days | 8 – 14 days | > 14 days |
| --- | --- | --- | --- |
| **LFA positive results** |  |  |  |
| *Alltest (Alltest)* | 5/18<br>(27.8) | 9/21<br>(42.9) | 35/41<br>(85.4) |
| *One Step (Innovita)* | 5/18<br>(27.8) | 14/21<br>(66.7) | 40/41<br>(97.6) |
| *Seroflash (Epigentek)* | 4/18<br>(22.2) | 11/21<br>(52.4) | 39/41<br>(95.1) |
| **ELISA positive results** |  |  |  |
| *Dia.Pro (Dia.Pro)* | 9/16*<br>(56.3) | 18/21<br>(85.7) | 40/41<br>(97.6) |
| **CLIA positive results\*\*** |  |  |  |
| *COV2T (Siemens)* | 2/5<br>(40.0) | 3/4<br>(75.0) | 41/41<br>(100.0) |
| *Elecsys (Roche)* | 2/5<br>(40.0) | 2/4<br>(50.0) | 40/41<br>(97.6) |

**Statistics:** values are expressed as absolute count (percentage). A positive serologic result was defined for LFA and ELISA tests for samples that resulted positive for either IgM or IgG antibodies. **Abbreviations:** LFA: lateral flow assay; ELISA: enzyme-linked immunoassay; CLIA: chemiluminescence.
\* Two samples presented indeterminate result for IgG or IgM and were excluded from the analysis
\*\* Sensitivity evaluation of CLIA techniques could only be performed with 50 samples due to insufficient sample volume

**Table 3.** Agreement between serologic assays

| Comparisons between serologic techniques | Results for IgM |  | Results for IgG |  | Overall (IgM/IgG) |  |
| --- | --- | --- | --- | --- | --- | --- |
|  | Agreement (%) | Kappa | Agreement (%) | Kappa | Agreement (%) | Kappa |
| **LFAs** |  |  |  |  |  |  |
| *Alltest vs. One Step* | 80.7 | 0.511 | 86.4 | 0.691 | 87.1 | 0.730 |
| *Alltest vs. SeroFlash* | 73.6 | 0.376 | 88.6 | 0.738 | 85.0 | 0.681 |
| *One Step vs. SeroFlash* | 91.4 | **0.815** | 90.7 | 0.788 | 92.1 | **0.838** |
| **LFAs and ELISA\*** |  |  |  |  |  |  |
| *Alltest vs. Dia.Pro* | 75.0 | 0.440 | 81.9 | 0.630 | 83.3 | 0.664 |
| *One Step vs. Dia.Pro* | 91.9 | **0.830** | 86.2 | 0.719 | 94.2 | **0.884** |
| *SeroFlash vs. Dia.Pro* | 89.7 | 0.785 | 85.5 | 0.704 | 88.4 | 0.767 |
| **LFAs and CLIAs\*\*** |  |  |  |  |  |  |
| *Alltest vs. COV2T* | - | - | - | - | 93.6 | **0.866** |
| *One Step vs. COV2T* | - | - | - | - | 99.1 | **0.981** |
| *SeroFlash vs. COV2T* | - | - | - | - | 95.5 | **0.906** |
| *Alltest vs. Elecsys* | - | - | - | - | 93.6 | **0.865** |
| *One Step vs. Elecsys* | - | - | - | - | 97.3 | **0.943** |
| *SeroFlash vs. Elecsys* | - | - | - | - | 93.6 | **0.868** |
| **ELISA and CLIAs** |  |  |  |  |  |  |
| *Dia.Pro vs. COV2T* | - | - | - | - | 98.2 | **0.963** |
| *Dia.Pro vs. Elecsys* | - | - | - | - | 96.4 | **0.925** |
| **CLIAs** |  |  |  |  |  |  |
| *COV2T vs. Elecsys* | - | - | - | - | 98.2 | **0.962** |

**Statistics:** values are expressed as percentage. A positive serologic result was defined for LFA and ELISA tests for samples that resulted positive for either IgM or IgG antibodies. Since CLIA techniques evaluate the presence of total antibodies (IgM+IgG), their agreement was only evaluated for the overall result of the test. Kappa scores over 0.81 (almost perfect agreement) are shown in bold.
**Abbreviations:** LFA: lateral flow assay; ELISA: enzyme-linked immunoassay; CLIA: chemiluminescence.
\* Two samples presented indeterminate result for IgG or IgM and were excluded from the analysis
\*\* Evaluation of agreement with CLIA techniques could only be performed with 50 samples due to insufficient sample volume

## 4. Discussion

Our study shows that *One Step*, *Dia.Pro*, *Elecsys* and *COV2T* achieved the best diagnostic performance results. All these techniques showed a specificity of 100% and sensitivities over 97% from 14 days after the onset of symptoms, as well as excellent levels of agreement between them.

Serologic tests have emerged as complementary tools to PCR in the diagnosis of SARS-CoV-2, including subclinical presentations [10]. Consequently, an increasing number of commercial tests have been developed. **Table 4** contains a summary of the studies that have evaluated different commercial immunoassays. Regarding LFAs, *NG Biotech* [11], *LabOn Time*, *Avioq* and *QuickZen* [12], showed similar results with *One Step* and *SeroFlash* in terms of sensitivity and specificity. Regarding ELISAs, *Euroimmun* has been the most frequently evaluated one, presenting overall sensitivities of 71.1 – 87.4% [11–14] and sensitivities from 14 days after the onset of symptoms ranging from 93.9 to 100%. However, Nicol *et al*. [11] and Montesinos *et al*. [12] reported lower specificity results (82.0 – 87.5%) for this ELISA than other authors [13,14]. Regarding CLIAs, *Abbott* showed sensitivities from 14 days ranging from 84.2 to 100% [11,15] and a high specificity (94.6 – 100%) [11,13,15]. Finally, in the two studies that evaluated the diagnostic performance of *Elecsys*, the reported sensitivities and specificities were lower than those obtained in our study [13,16]. In our experience, *Dia.Pro*, *Elecsys* and *COV2T* obtained excellent levels of sensitivity, specificity and agreement. We also showed that those LFAs that use recombinant nucleocapsid and spike antigens (*One Step* and *SeroFlash*) could achieve practically the same diagnostic performance results than ELISA and CLIAs. At the beginning of its development, the usefulness of LFAs was questioned due to a lack of official performance validations [5,17]. Nowadays these tests have demonstrated their usefulness and reliability in epidemiological studies [7], and also in the diagnosis of pneumonia of unknown etiology with negative PCR for SARS-CoV-2 [18].

**Table 4.**
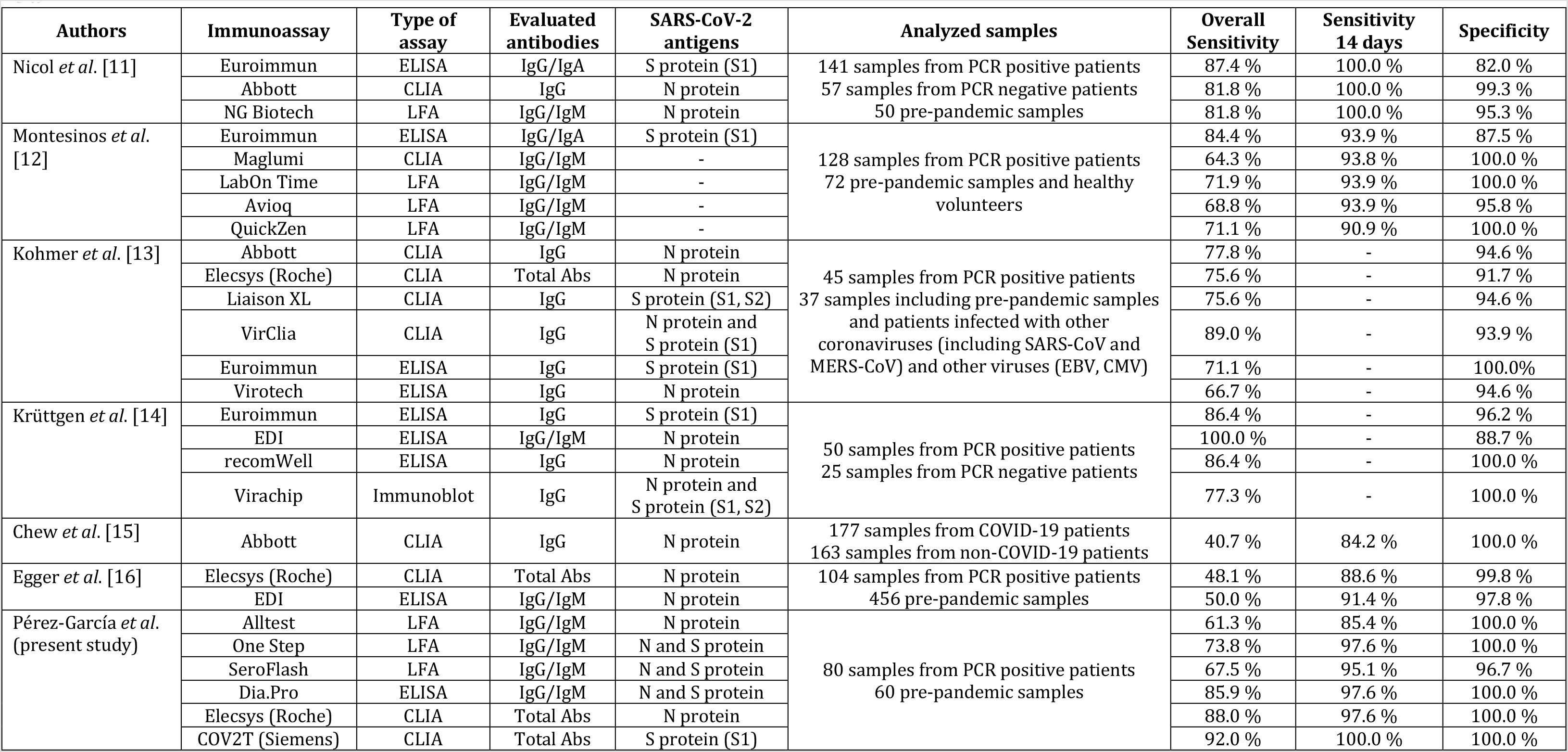
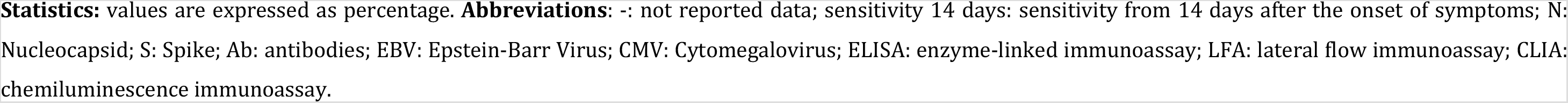
Studies that have evaluated commercial immunoassays to detect antibodies against SARS-CoV-2

| Authors | Immunoassay | Type of assay | Evaluated antibodies | SARS-CoV-2 antigens | Analyzed samples | Overall Sensitivity | Sensitivity 14 days | Specificity |
| --- | --- | --- | --- | --- | --- | --- | --- | --- |
| Nicol *et al.* [11] | Euroimmun | ELISA | IgG/IgA | S protein (S1) | 141 samples from PCR positive patients<br>57 samples from PCR negative patients<br>50 pre-pandemic samples | 87.4 % | 100.0 % | 82.0 % |
|  | Abbott | CLIA | IgG | N protein |  | 81.8 % | 100.0 % | 99.3 % |
|  | NG Biotech | LFA | IgG/IgM | N protein |  | 81.8 % | 100.0 % | 95.3 % |
| Montesinos *et al.* [12] | Euroimmun | ELISA | IgG/IgA | S protein (S1) | 128 samples from PCR positive patients<br>72 pre-pandemic samples and healthy volunteers | 84.4 % | 93.9 % | 87.5 % |
|  | Maglumi | CLIA | IgG/IgM | - |  | 64.3 % | 93.8 % | 100.0 % |
|  | LabOn Time | LFA | IgG/IgM | - |  | 71.9 % | 93.9 % | 100.0 % |
|  | Avioq | LFA | IgG/IgM | - |  | 68.8 % | 93.9 % | 95.8 % |
|  | QuickZen | LFA | IgG/IgM | - |  | 71.1 % | 90.9 % | 100.0 % |
| Kohmer *et al.* [13] | Abbott | CLIA | IgG | N protein | 45 samples from PCR positive patients<br>37 samples including pre-pandemic samples and patients infected with other coronaviruses (including SARS-CoV and MERS-CoV) and other viruses (EBV, CMV) | 77.8 % | - | 94.6 % |
|  | Elecsys (Roche) | CLIA | Total Abs | N protein |  | 75.6 % | - | 91.7 % |
|  | Liaison XL | CLIA | IgG | S protein (S1, S2) |  | 75.6 % | - | 94.6 % |
|  | VirClia | CLIA | IgG | N protein and S protein (S1) |  | 89.0 % | - | 93.9 % |
|  | Euroimmun | ELISA | IgG | S protein (S1) |  | 71.1 % | - | 100.0 % |
|  | Virotech | ELISA | IgG | N protein |  | 66.7 % | - | 94.6 % |
| Krüttgen *et al.* [14] | Euroimmun | ELISA | IgG | S protein (S1) | 50 samples from PCR positive patients<br>25 samples from PCR negative patients | 86.4 % | - | 96.2 % |
|  | EDI | ELISA | IgG/IgM | N protein |  | 100.0 % | - | 88.7 % |
|  | recomWell | ELISA | IgG | N protein |  | 86.4 % | - | 100.0 % |
|  | Virachip | Immunoblot | IgG | N protein and S protein (S1, S2) |  | 77.3 % | - | 100.0 % |
| Chew *et al.* [15] | Abbott | CLIA | IgG | N protein | 177 samples from COVID-19 patients<br>163 samples from non-COVID-19 patients | 40.7 % | 84.2 % | 100.0 % |
| Egger *et al.* [16] | Elecsys (Roche) | CLIA | Total Abs | N protein | 104 samples from PCR positive patients<br>456 pre-pandemic samples | 48.1 % | 88.6 % | 99.8 % |
|  | EDI | ELISA | IgG/IgM | N protein |  | 50.0 % | 91.4 % | 97.8 % |
| Pérez-García *et al.* (present study) | Alltest | LFA | IgG/IgM | N protein | 80 samples from PCR positive patients<br>60 pre-pandemic samples | 61.3 % | 85.4 % | 100.0 % |
|  | One Step | LFA | IgG/IgM | N and S protein |  | 73.8 % | 97.6 % | 100.0 % |
|  | SeroFlash | LFA | IgG/IgM | N and S protein |  | 67.5 % | 95.1 % | 96.7 % |
|  | Dia.Pro | ELISA | IgG/IgM | N and S protein |  | 85.9 % | 97.6 % | 100.0 % |
|  | Elecsys (Roche) | CLIA | Total Abs | N protein |  | 88.0 % | 97.6 % | 100.0 % |
|  | COV2T (Siemens) | CLIA | Total Abs | S protein (S1) |  | 92.0 % | 100.0 % | 100.0 % |

**Statistics:** values are expressed as percentage. **Abbreviations:** -: not reported data; sensitivity 14 days: sensitivity from 14 days after the onset of symptoms; N: Nucleocapsid; S: Spike; Ab: antibodies; EBV: Epstein-Barr Virus; CMV: Cytomegalovirus; ELISA: enzyme-linked immunoassay; LFA: lateral flow immunoassay; CLIA: chemiluminescence immunoassay.

Our study presents some limitations. First, it is a retrospective study that has been conducted in a single institution. Further prospective multicenter studies are necessary to reinforce our findings. Second, sensitivity evaluation of CLIAs was performed only over 50 sera, due to insufficient simple volume. However, the samples that could not be analyzed belonged to the first two weeks after the onset of symptoms. As a consequence, this limitation did not affect the results about sensitivity from 14 days, when the vast majority of patients seroconvert according to different studies [10,12,18]. Finally, we have analyzed the results of six among all commercialized immunoassays. Consequently, our results should not be extrapolated to other available immunoassays and more comparative studies and meta-analysis are needed to establish the usefulness of other serologic tests.

In conclusion, *One Step* LFA, *Dia.Pro* ELISA and *Elecsys* and *COV2T CLIAs* present the best diagnostic performance results. All these techniques showed a specificity of 100% and sensitivities over 97% from 14 days after the onset of symptoms, as well as excellent levels of agreement between them. To our knowledge, this study constitutes the first comparative evaluation of these six immunoassays. These findings indicate that these tests could be reliable tools for the diagnosis of COVID-19 and the performance of epidemiological studies.

## Data Availability

The datasets used and analyzed during the current study may be available from the corresponding author upon reasonable request.

## Acknowledgements

We thank Rebeca Baileń for her help with the preparation of the manuscript.

## Compliance with Ethical Standards

### Informed consent

Since the present study is retrospective, informed consent was not required.

### Ethical approval

The study was conducted according to the ethical requirements established by the Declaration of Helsinki. The Ethics Committee of Hospital Universitario Príncipe de Asturias (Madrid) approved the study (protocol code: Comparativa Sero-COVID).

## Author contributions

Study concept and design: FPG and JCG

Patients’ selection and clinical data acquisition: FPG, RPT, MEI, JR, TA, PGH and RG

Sample processing: JR, RPT, MEI, TA, RG and PGH

Statistical analysis and interpretation of data: FPG

Writing of the manuscript: FPG and JCG

Critical revision of the manuscript for relevant intellectual content: JCG

Supervision and visualization: JCG

All authors read and approved the final manuscript.

